# Multimodal AI-driven Biomarker for Early Detection of Cancer Cachexia

**DOI:** 10.1101/2025.09.17.25336028

**Authors:** Sabeen Ahmed, Nathan Parker, Margaret Park, Evan W. Davis, Daniel Jeong, Jennifer B. Permuth, Matthew B. Schabath, Yasin Yilmaz, Ghulam Rasool

## Abstract

Cancer cachexia, a multifactorial metabolic syndrome characterized by severe muscle wasting and weight loss, contributes to poor outcomes across various cancer types but lacks a standardized, generalizable biomarker for early detection. We present a multimodal AI-based biomarker trained on real-world clinical, radiologic, laboratory, and unstructured clinical note data, leveraging foundation models and large language models (LLMs) to identify cachexia at the time of cancer diagnosis. Prediction accuracy improved with each added modality: 77% using clinical variables alone, 81% with added laboratory data, and 85% with structured symptom features extracted from clinical notes. Incorporating embeddings from clinical text and CT images further improved accuracy to 92%. The framework also demonstrated prognostic utility, improving survival prediction as data modalities were integrated. Designed for real-world clinical deployment, the framework accommodates missing modalities without requiring imputation or case exclusion, supporting scalability across diverse oncology settings. Unlike prior models trained on curated datasets, our approach utilizes standard-of-care clinical data, facilitating integration into oncology workflows. In contrast to fixed-threshold composite indices such as the cachexia index (CXI), the model generates patient-specific predictions, enabling adaptable, cancer-agnostic performance. To enhance clinical reliability and safety, the framework incorporates uncertainty estimation to flag low-confidence cases for expert review. This work advances a clinically applicable, scalable, and trustworthy AI-driven decision support tool for early cachexia detection and personalized oncology care.

## 1. Introduction

Cancer cachexia is a complex, multifactorial metabolic syndrome strongly associated with poor quality of life and reduced survival. Prevalence of cancer cachexia varies by cancer type, reaching 60–70% in gastroesophageal and pancreatic cancers and 40–50% in hematological, colorectal, and lung cancers [1]. The key drivers, chronic inflammation, metabolic disturbances, hormonal shifts, and reduced food intake, are commonly assessed using indicators like elevated C-reactive protein (CRP) and low serum albumin [2, 3]. However, these indicators are not specific to cancer cachexia and can be influenced by other conditions, limiting their diagnostic utility. Radiological skeletal muscle metrics, including skeletal muscle area (SMA) and skeletal muscle index (SMI), provide additional insight but can be confounded by treatment-induced fluctuations. Composite indices such as the cachexia index (CXI) [4, 5], modified CXI (mCXI) [6], and cancer cachexia risk score (CCRS) [7] combine SMI with nutritional and inflammatory markers. However, these indices rely on fixed threshold-based cutoffs that fail to capture inter-patient variability, reducing their generalizability across diverse cancer populations. Tools like CASCO [8], while comprehensive, are resource-intensive and unsuitable for routine clinical use. Additionally, current approaches typically assess cachexia after clinically apparent muscle wasting or weight loss has occurred, limiting opportunities for early intervention. Fragmentation of data across modalities (clinical, laboratory, imaging, and unstructured text) further complicates early detection efforts. Variability in documentation practices, imaging protocols, and biomarker thresholds across institutions poses further challenges to standardizing cachexia assessment in real-world settings. These limitations underscore the need for a standardized, scalable, and adaptable approach for early cancer cachexia detection.

To address these limitations, we propose a multimodal AI biomarker that integrates routinely collected clinical data (including patient demographics, anthropometric measurements, and disease status), laboratory values, radiologic metrics, and unstructured clinical notes to enable early detection of cancer cachexia, with a design focused on real-world clinical adaptation [9–12]. By leveraging data acquired during standard-of-care cancer management, the model supports generalizability, scalability, and seamless integration into routine oncology workflows [13]. Importantly, the framework is engineered to handle missing modalities without compromising predictive performance, enhancing its applicability across diverse clinical settings where data completeness may vary. We incorporate uncertainty estimation to support self-assessment, enabling the model to flag low-confidence cases for expert review and ensuring that clinical decision-making remains both data-driven and safe [14–16]. Unlike traditional composite indices constrained by static thresholds, our adaptive framework generates patient-specific predictions based on individual characteristics (e.g., age, ethnicity, cancer type, and stage), promoting fairness and mitigating bias. This aligns with the current priorities in multimodal AI research, which emphasize the development of clinically deployable, robust, and ethically sound AI tools for oncology care.

Our results demonstrate that integrating multiple data modalities substantially improves cachexia prediction accuracy at the time of diagnosis. In addition to early detection, survival analysis showed significantly improved prognostic performance as additional modalities were incorporated. Together, these findings highlight the potential of the proposed multimodal AI biomarker to enable timely interventions, personalize treatment strategies, and improve outcomes for patients at risk of cancer cachexia. **Figure 1** illustrates the proposed multimodal AI framework, which integrates clinical, laboratory, radiologic, and unstructured text data through a stepwise configuration strategy. The framework accommodates varying levels of data completeness, incorporates uncertainty-aware prediction to flag low-confidence cases, and supports clinical scalability through modular design and progressive data fusion.

**Figure 1:**
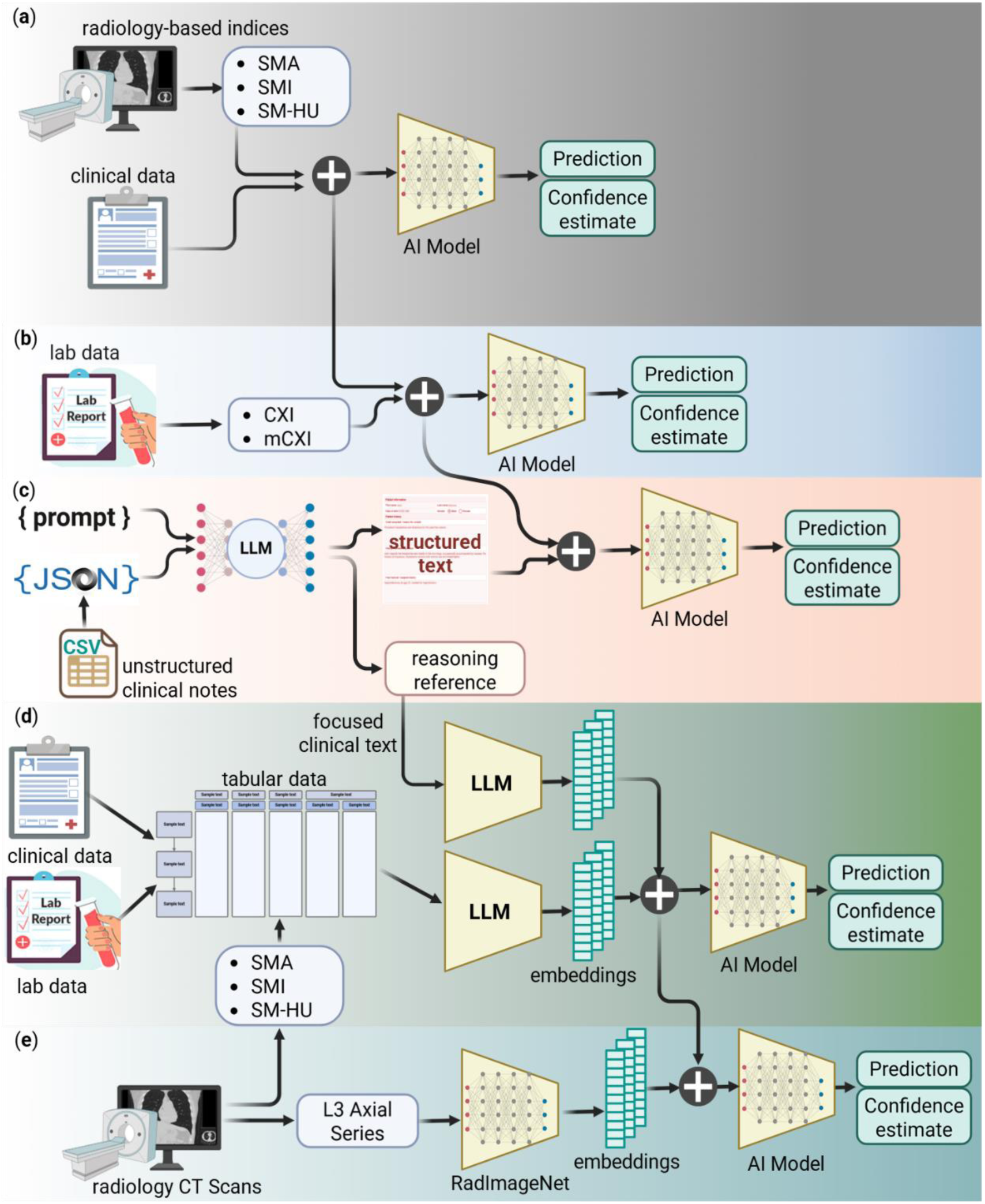
Multimodal AI Framework for Cancer Cachexia Prediction. Stepwise integration of data modalities was used to train AI models for cancer cachexia detection. The five primary configurations include: (**a**) clinical data and CT-derived skeletal muscle metrics, including skeletal muscle area (SMA), skeletal muscle index (SMI), and skeletal muscle radiodensity (SM-HU); (**b**) addition of laboratory data and derived cachexia-related indices, such as the cachexia index (CXI) and modified cachexia index (mCXI); (**c**) further inclusion of structured symptom features extracted from unstructured clinical notes using large language models (LLMs); (**d**) concatenation of tabular features in (b) with LLM-generated embeddings from symptom-focused clinical text; and (**e**) addition of CT image embeddings from the third lumbar (L3) vertebral level generated using a radiology foundation model (RadImageNet). A sixth experimental configuration integrated clinical, laboratory, and structured note features while deliberately excluding skeletal muscle metrics. The framework includes an uncertainty-aware prediction mechanism that flags low-confidence outputs for expert review, supporting reliable integration into clinical workflows.

The key contributions of this study include:

1. **Development of a multimodal AI framework** that integrates routinely collected patient data, including demographics, anthropometric measures, cancer staging, radiologic skeletal muscle metrics, laboratory biomarkers, and cachexia-related symptoms extracted from unstructured clinical notes, for early detection of cancer cachexia at the time of cancer diagnosis.
2. **Application of LLMs** to transform unstructured clinical notes into structured symptom features, enabling seamless incorporation of text-derived information into predictive pipelines and improving model performance.
3. **Implementation of multimodal fusion strategies**, combining embeddings generated from heterogeneous data modalities, including tabular clinical variables, radiologic images and derived metrics, laboratory data, and focused clinical text, using foundation models and LLMs to enhance cachexia detection accuracy.
4. **Extension of the framework to survival prediction**, demonstrating that stepwise integration of multimodal data not only improves early detection but also enhances prognostic modeling performance.
5. **Design of a clinically adaptable, uncertainty-aware AI framework**, capable of accommodating missing modalities, supporting human-in-the-loop decision-making, and facilitating integration into real-world oncology workflows to enable timely interventions and personalized care.

The Methods section describes the dataset, multimodal framework, model development, and evaluation approach in detail.

## 2. Materials and Methods

This retrospective cohort study was conducted using data from the Florida Pancreas Collaborative (FPC), a statewide, prospective multi-institutional study focused on patients with pancreatic ductal adenocarcinoma (PDAC) [17]. The final analysis included 236 PDAC patients with known SMA and cachexia status information, referred to as Cohort I. Of these, 131 patients were treated at the coordinating center, Moffitt Cancer Center, and had additional data accessible to our team in the EHR; they are referred to as Cohort II in this manuscript. **Figure 2** illustrates the branching of the overall cohort into Moffitt and non-Moffitt subsets and the data availability across modalities. **Table 1** summarizes key demographic and clinical characteristics for both cohorts. This research was approved by the Moffitt Cancer Center Scientific Review Committee and the Advarra Institutional Review Board.

**Figure 2:**
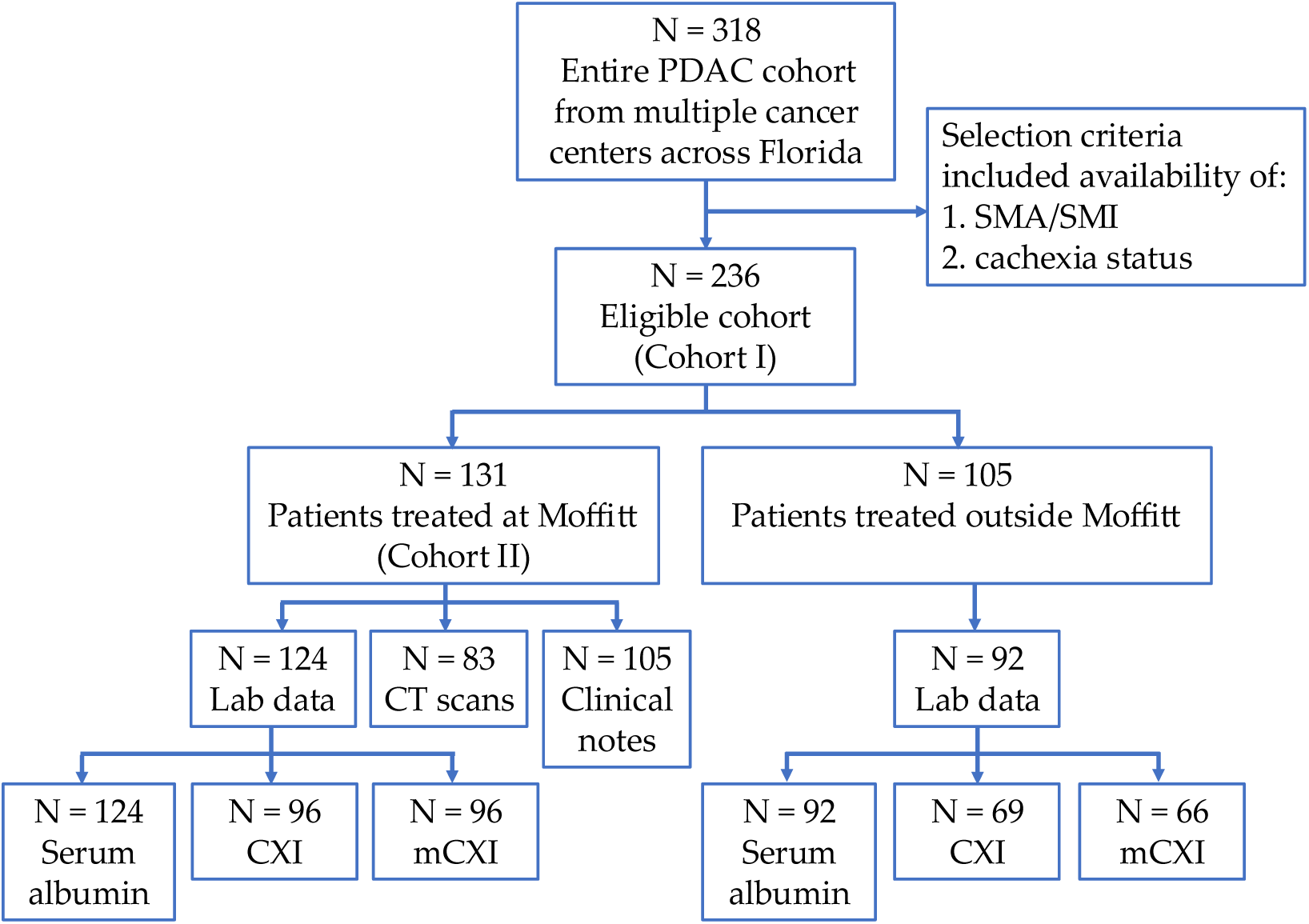
Cohort stratification by institution and data modality availability. Flow diagram illustrating the branching of the full PDAC cohort (Cohort I, *n* = 236) into Moffitt Cancer Center patients (Cohort II, *n* = 131) and non-Moffitt patients (*n* = 105). The availability of clinical notes, CT scans, and laboratory data within each subgroup is also shown, highlighting data completeness across modalities.

**Table 1.** – Demographic and clinical characteristics of the PDAC patient cohorts included in this study. Summary of patient characteristics for the full cohort (Cohort I, n = 236) and the Moffitt-only subset (Cohort II, n = 131).

|  | *Cohort I (n = 236)* | *Cohort II (n = 131)* |
| --- | --- | --- |
| Age at diagnosis, mean (SD) | 69.05 ± 10.13 | 69.57 ± 10.52 |
| Weight (lbs.) at diagnosis, mean (SD) | 166.95 ± 37.88 | 166.78 ± 36.37 |
| Height (m), mean (SD) | 1.69 ± 0.59 | 2.82 ± 0.35 |
| BMI at diagnosis, mean (SD) | 26.75 ± 5.77 | 26.91 ± 5.50 |
| Sex, N |  |  |
| Female | 119 | 65 |
| Male | 117 | 66 |
| Race and Ethnicity, N |  |  |
| Non-Hispanic White | 176 | 114 |
| Hispanic/Latinx | 36 | 10 |
| Non-Hispanic Black | 24 | 7 |
| TNM Stage (Pathological), N |  |  |
| 1: IA (T1, N0, M0) | 15 | 9 |
| 2: IB (T2, N0, M0) | 27 | 17 |
| 3: IIA (T3, N0, M0) | 26 | 18 |
| 4: IIA (T1, N1, M0) | 1 | 1 |
| 5: IIA (T2, N1, M0) | 17 | 8 |
| 6: IIB (T3, N1, M0) | 16 | 4 |
| 7: III (T4, Any N, M0) | 29 | 26 |
| 8: IV (Any T, Any N, M1) | 71 | 47 |
| 9: Tumor stage cannot be assessed | 13 | 0 |
| -1: NA | 21 | 1 |
| Cachexia Status, N |  |  |
| Cachectic | 152 | 79 |
| Non-cachectic | 84 | 52 |
| Patient Hospital, N |  |  |
| Moffitt | 131 | 131 |
| Outside Moffitt | 105 | 0 |
| Model Training and Evaluation, N |  |  |
| Training set (cross-validation) | 210 | 105 |
| Held-out test set | 26 | 26 |

### 2.1. Data Sources and Modalities

We integrated the following four primary data modalities available at the time of cancer diagnosis and formed incremental combinations of these modalities, given in **Table 3**:

1. **Clinical data**: Including demographics (sex, race, ethnicity, and age), anthropometric measurements (weight, height, and BMI), and cancer staging (TNM), summarized in **Table 1**.
2. **Laboratory data**: Capturing nutritional and inflammatory biomarkers and derived indices indicating cachexia status listed in **Table 2**.
3. **Radiologic data**: Consisting of skeletal muscle metrics (SMA, SMI, SM-HU) derived from CT imaging at the third lumbar vertebra (L3), and embeddings generated from these L3 CT images.
4. **Unstructured clinical notes**: Extracted from electronic health record (EHR) and processed using LLMs. We used the following types of notes: (i) nutrition assessment form, (ii) nutrition diagnosis/comments, (iii) progress note, (iv) dietary assessment, evaluation, or comments, (v) inter-visit note, (vi) ambulatory care note, (vii) history and physical note, and (viii) patient assessment.

**Table 2:**
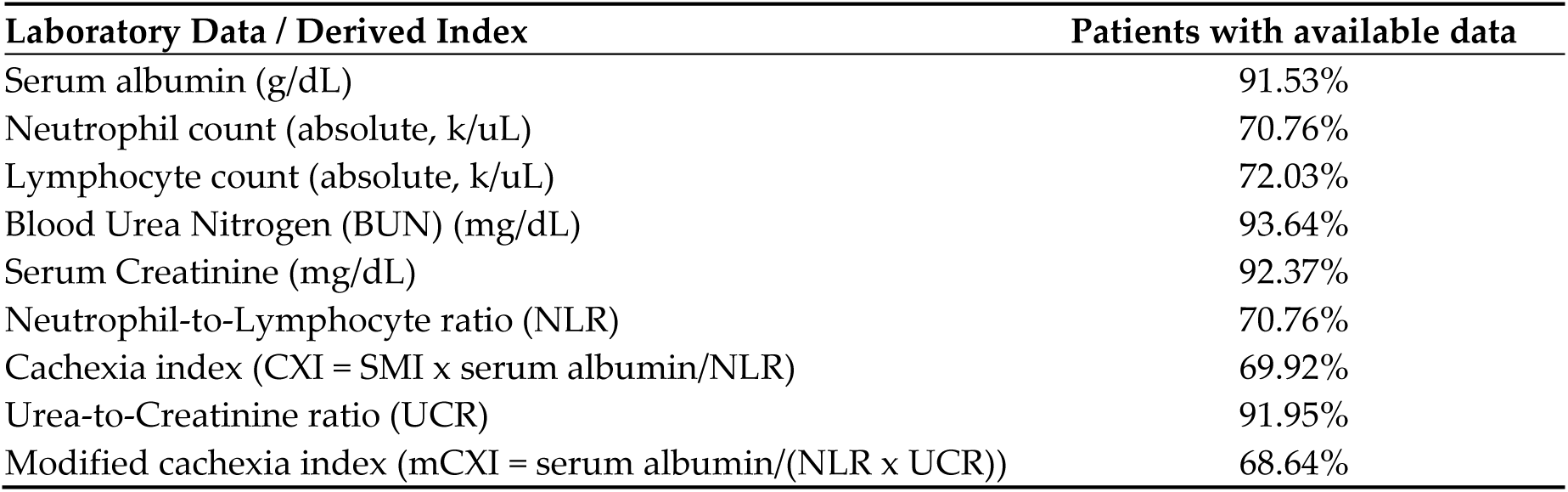
Laboratory biomarkers and derived cachexia-related indices used in this study (Cohort I, n=236)

**Table 3:**
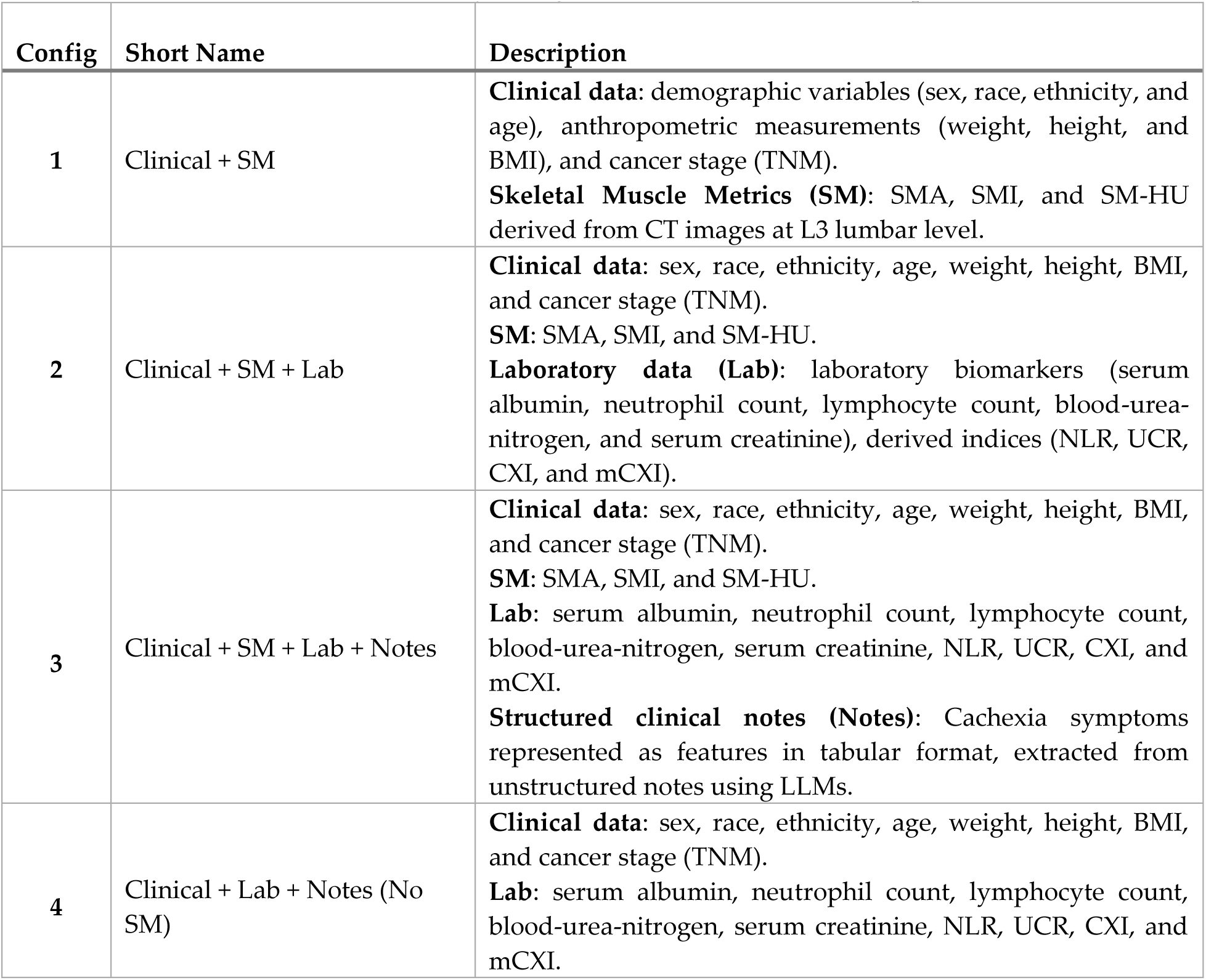
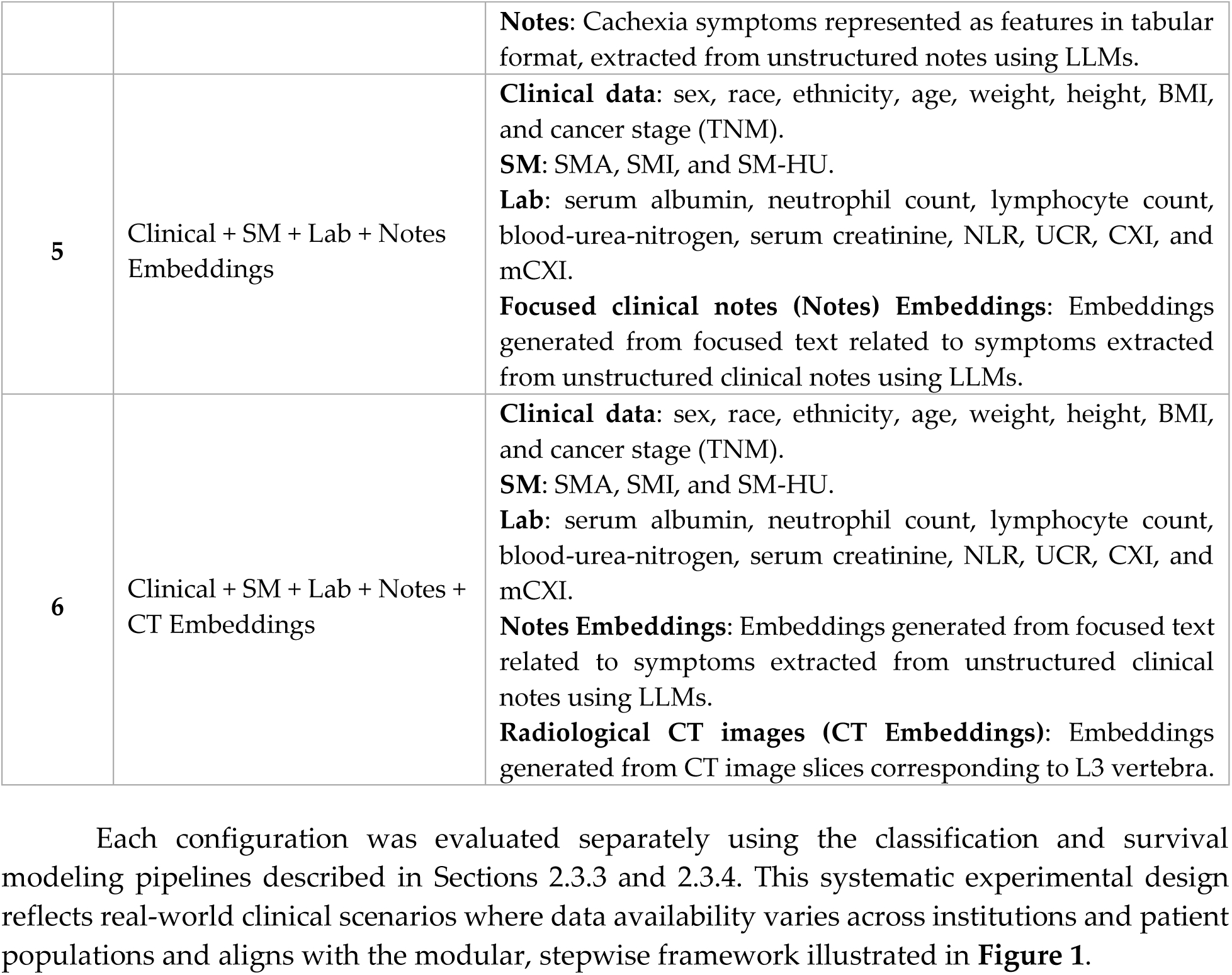
Definition of the six data modality configurations used in model development.

| Config | Short Name | Description |
| --- | --- | --- |
| 1 | Clinical + SM | **Clinical data:** demographic variables (sex, race, ethnicity, and age), anthropometric measurements (weight, height, and BMI), and cancer stage (TNM).<br>**Skeletal Muscle Metrics (SM):** SMA, SMI, and SM-HU derived from CT images at L3 lumbar level. |
| 2 | Clinical + SM + Lab | **Clinical data:** sex, race, ethnicity, age, weight, height, BMI, and cancer stage (TNM).<br>**SM:** SMA, SMI, and SM-HU.<br>**Laboratory data (Lab):** laboratory biomarkers (serum albumin, neutrophil count, lymphocyte count, blood-urea-nitrogen, and serum creatinine), derived indices (NLR, UCR, CXI, and mCXI). |
| 3 | Clinical + SM + Lab + Notes | **Clinical data:** sex, race, ethnicity, age, weight, height, BMI, and cancer stage (TNM).<br>**SM:** SMA, SMI, and SM-HU.<br>**Lab:** serum albumin, neutrophil count, lymphocyte count, blood-urea-nitrogen, serum creatinine, NLR, UCR, CXI, and mCXI.<br>**Structured clinical notes (Notes):** Cachexia symptoms represented as features in tabular format, extracted from unstructured notes using LLMs. |
| 4 | Clinical + Lab + Notes (No SM) | **Clinical data:** sex, race, ethnicity, age, weight, height, BMI, and cancer stage (TNM).<br>**Lab:** serum albumin, neutrophil count, lymphocyte count, blood-urea-nitrogen, serum creatinine, NLR, UCR, CXI, and mCXI. |
|  |  | **Notes:** Cachexia symptoms represented as features in tabular format, extracted from unstructured notes using LLMs. |
| 5 | Clinical + SM + Lab + Notes Embeddings | **Clinical data:** sex, race, ethnicity, age, weight, height, BMI, and cancer stage (TNM).<br>**SM:** SMA, SMI, and SM-HU.<br>**Lab:** serum albumin, neutrophil count, lymphocyte count, blood-urea-nitrogen, serum creatinine, NLR, UCR, CXI, and mCXI.<br>**Focused clinical notes (Notes) Embeddings:** Embeddings generated from focused text related to symptoms extracted from unstructured clinical notes using LLMs. |
| 6 | Clinical + SM + Lab + Notes + CT Embeddings | **Clinical data:** sex, race, ethnicity, age, weight, height, BMI, and cancer stage (TNM).<br>**SM:** SMA, SMI, and SM-HU.<br>**Lab:** serum albumin, neutrophil count, lymphocyte count, blood-urea-nitrogen, serum creatinine, NLR, UCR, CXI, and mCXI.<br>**Notes Embeddings:** Embeddings generated from focused text related to symptoms extracted from unstructured clinical notes using LLMs.<br>**Radiological CT images (CT Embeddings):** Embeddings generated from CT image slices corresponding to L3 vertebra. |

### 2.2. Data Processing

#### 2.2.1. Clinical Data

The clinical data included demographic variables (age, sex, race, and ethnicity), anthropometric measurements (weight, height, and BMI), and cancer staging information. To address missing data, age values were imputed using the mean age of the entire study population. For weight and height, missing values were imputed using sex– and race/ethnicity-specific means to preserve underlying population distributions. Tumor stage was determined using the Tumor, Node, Metastasis (TNM) classification system. When not available or not assessable, the tumor stage was coded as ‘-1’ to represent missingness. All categorical variables, including sex, race/ethnicity, and tumor stage, were binarized before being used in machine learning models. Patients were categorized into cancer cachexia stages to generate ground truth labels for training and evaluation of the deep learning models, using criteria defined in the literature. The two-stage system introduced by Fearon et al. [3] categorizes patients as either cachectic or non-cachectic. A patient was termed cachectic if either of the following conditions were met:

a. Weight loss was >5% over the past six months for BMI ≥ 20.
b. Weight loss was >2% over the past six months for a BMI < 20.

For the embedding-based experiments, structured clinical variables in tabular format were transformed into a text-based format suitable for large language model input. The GatorTron-medium model [19], pretrained on EHR text, was used to generate clinical data embeddings. Where applicable, the word “missing” was explicitly inserted to preserve information on missingness. For qualitative variables, the original text descriptors were retained. This conversion enabled the effective integration of tabular clinical data into the multimodal deep learning pipeline alongside laboratory, radiologic, and unstructured text features.

#### 2.2.2. Laboratory Data

Laboratory data included a panel of routinely collected serum biomarkers that reflect nutritional and inflammatory status at the time of cancer diagnosis. Specifically, the following laboratory values were extracted for each patient: serum albumin (g/dL), absolute neutrophil count (k/uL), absolute lymphocyte count (k/uL), blood urea nitrogen (BUN; mg/dL), and serum creatinine (mg/dL). **Table 2** and **Figure 2** summarize the availability of the laboratory data and the derived indices used in this study, across the cohorts.

To capture established biomarkers associated with cancer cachexia, we derived several composite indices from the laboratory data. The neutrophil-to-lymphocyte ratio (NLR) was calculated by dividing the absolute neutrophil count by the absolute lymphocyte count. The cachexia index (CXI), as defined by Jafri et al. [4], was computed using the formula:

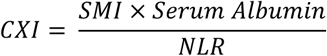

The modified cancer cachexia index (mCXI), introduced by Yuan et al. [6], was calculated using the following formula:

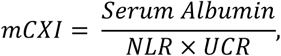

where UCR represents the urea-to-creatinine ratio, calculated as BUN divided by serum creatinine. The missing laboratory values were handled using a consistent placeholder strategy. If a specific laboratory result was not available for a patient, the corresponding value was set to ‘-’ to signify missingness. Similarly for derived indices such as NLR, CXL, mCXL and UCR, missing inputs Variables propagated to missing index values, which were likewise coded as ‘-1’. This strategy ensured that missingness was explicitly represented during model training and inference.

#### 2.2.3. Unstructured Clinical Notes

Unstructured clinical notes from EHR were used as an additional data modality to capture contextual information on patient symptoms, functional status, and nutritional assessments at the time of cancer diagnosis. To leverage these notes for machine learning, we processed them using two complementary ways: structured feature extraction and text embedding generation.

1. **Structured Feature Extraction**: All available notes for each patient were combined in a JSON file. These textual data notes were processed using Llama3.2:3b-instruct-fp16, Qwen2.5:7b-instruct-fp16, and DeepSeek-r1:70b LLMs running on local GPU machines served by Ollama [20–22]. The LLM was used to extract responses in “yes”, “no”, and “not-given” format based on the notes available for each patient, for a set of questions relevant to determining cancer cachexia status. The LLM was instructed to provide reasoning for its response along with references from the text. The response to these questions was converted into a tabular format, with ‘1’ representing ‘yes’, ‘0’ representing ‘no’, and ‘-1’ representing ‘not-given.’ This process generated a structured, patient-level symptom feature set suitable for inclusion in the multimodal machine learning models.
2. **Text Embedding Generation**: In parallel, we generated semantic embeddings from the unstructured notes to capture latent textual patterns that could enhance model performance. Rather than embedding the full raw text, we focused on the reasoning and reference text sections extracted by the LLMs for each patient, reducing noise and maximizing clinical relevance. These extracted text segments were processed using the GatorTron-medium model [19], a large language model pretrained on electronic health record data. To accommodate the Model’s 512-tpken input limit, each patient’s text was segmented into 512-token chunks, and embeddings were generated for each chunk. The final patient-level embedding was obtained by applying average pooling across all chunks.

Together, these structured features and textual embeddings allowed the model to incorporate both discrete symptom signals and richer linguistic representations from clinical notes, contributing to improved cachexia detection performance.

#### 2.2.4. Radiologic Data

Radiological data focused on skeletal muscle metrics and imaging-based embeddings derived from abdominal CT scans obtained at the time of cancer diagnosis. CT scans were only available for patients treated at Moffitt Cancer Center (Cohort II). SMA, SMI, and SM-HU were quantified from axial CT slices at the third lumbar vertebra (L3) level. For Moffitt patients, these metrics were obtained using SMAART-AI [23], an automated pipeline that segments skeletal muscle and calculates the average muscle density. For patients from outside institutions, muscle measurements and metrics were extracted by a board-certified radiologist using the AW Server platform [24]. When multiple imaging phases were available, the venous phase axial series was prioritized for analysis, followed by the arterial phase if venous images were unavailable. SMI was calculated by dividing the SMA (cm^2^) by the square of height (m^2^).

For imaging-based deep learning experiments, we generated CT image embeddings to capture radiomic and anatomical features beyond skeletal muscle metrics. Using the HoNEYBEE platform [25] L3-level CT slices were processed through RadImageNet, a foundation model pretrained on radiology images [26]. All axial slices corresponding to the L3 level from each patient’s CT scan were used in this process. When multiple series were available for a patient, priority was given to the venous phase. Each CT slice was embedded separately, and average pooling was applied across slices to produce a single, patient-level imaging embedding. These radiologic features, both skeletal muscle indices and CT image embeddings, were integrated as distinct input modalities within the multimodal AI framework for cachexia detection and survival prediction.

### 2.3. Multimodal Data Integration and Experimental Design

To systematically evaluate the contribution of each data modality and assess the overall utility of a multimodal AI approach, we designed a two-part framework encompassing multimodal data integration and stepwise experimental configurations. First, we developed a flexible data fusion strategy capable of handling heterogeneous clinical, laboratory, radiologic, and unstructured text data. Second, we designed a series of experiments to measure the additive value of each modality and to compare performance across different input configurations (**Table 3**). This design reflects real-world clinical scenarios where data availability varies by patient and institution, and where scalable, generalizable models must operate effectively even in the presence of missing modalities.

#### 2.3.1. Multimodal Data Fusion Strategy

To integrate diverse patient data sources into a unified modeling pipeline, we implemented a multimodal data fusion strategy capable of combining structured, unstructured, and imaging-derived information. This fusion approach was designed to accommodate varying levels of data completeness across patients while maximizing the predictive value of each modality. For each patient, modality-specific feature representations were first generated during data preprocessing (Section 2.2). These included tabular features from clinical and laboratory data, structured symptom features extracted from unstructured clinical notes using LLMs, and embedding vectors from both focused clinical text and L3-level CT images.

For experiments based on structured features, we concatenated tabular clinical variables (demographics, anthropometrics, cancer stage), laboratory data (e.g., serum albumin, NLR, CXI, and mCXI), and skeletal muscle metrics (SMA, SMI, and SM-HU) to LLM-extracted structured note features into a single unified feature vector per patient. For embedding-based experiments, we concatenated GatorTron-based embeddings from focused clinical text and RadImageNet-based embeddings from L3 CT images to the feature vector containing clinical variables, laboratory-derived indices, and skeletal muscle metrics.

To tackle the challenge of missing data and modality (**Figure 2** and **Table 2**), we developed a comprehensive strategy that preserved input dimensionality across patients. For any missing modality, we inserted a predefined placeholder vector with fixed dimensions to represent missingness explicitly. This allowed the model to remain robust and generalizable across patients with incomplete data. This multimodal data fusion process served as the foundation for both classification and survival analysis tasks. The stepwise inclusion of modalities, as depicted in **Figure 1** enabled systematic evaluation of how each data source contributed to overall model performance.

#### 2.3.2. Experimental Configurations for Model Training

To systematically evaluate the incremental contribution of each data modality, we designed a series of experiments using stepwise configurations that progressively incorporated additional data sources. This approach allowed us to assess the additive value of laboratory data, structured features from unstructured clinical notes, and imaging-derived embeddings on model performance for cancer cachexia detection. **Table 3Table 3**: Definition of the six data modality configurations used in model development. provides a summary of the six data modality configurations used in this study.

Each configuration was evaluated separately using the classification and survival modeling pipelines described in Sections 2.3.3 and 2.3.4. This systematic experimental design reflects real-world clinical scenarios where data availability varies across institutions and patient populations and aligns with the modular, stepwise framework illustrated in **Figure 1**.

#### 2.3.3. Cancer Cachexia Classification

We framed cancer cachexia detection as a binary classification task. A multilayer perceptron (MLP) architecture was used for all classification experiments, consisting of four hidden layers with dropout regularization to mitigate overfitting. Key hyperparameters, including the number of nodes per layer, dropout rate, learning rate, and random seed, were optimized using a Bayesian search strategy implemented through the Weights & Biases (wandb) platform [27]. The use of random seeds ensured reproducibility across all experimental runs.

To evaluate the model’s robustness and generalizability, we employed cross-validation on training sets and evaluated the final model performance on a held-out test set. For the full cohort (Cohort I, *n* = 236), models were trained using 10-fold cross-validation on 210 patients, while the remaining 26 patients were held out for independent testing. Similarly, for the Moffitt-only subset (Cohort II, n = 131), 7-fold cross-validation was conducted using 105 patients, with 26 patients held out as a test set. For each experimental configuration shown in **Table 3**, five distinct MLP models were trained with different hidden layer node counts selected during hyperparameter optimization. Within each model, predictions were averaged across all cross-validation folds to obtain patient-level outputs. The final prediction for each patient in the test set was then obtained by averaging the predictions from the five cross-validated models, forming an ensemble that minimized variance and improved robustness. In addition to producing classification probabilities, we calculated the variance across ensemble outputs as a measure of predictive uncertainty [14], enabling identification of low-confidence predictions for expert review in a real-world clinical deployment scenario (**Figure 1**).

#### 2.3.4. Survival Analysis

In addition to classifying cachexia, we evaluated the prognostic value of multimodal features by predicting survival after PDAC diagnosis. Survival time was defined as the interval between diagnosis and either death or last follow-up. MLP models were trained using the Cox proportional hazards loss function, a standard method for modeling right-censored time-to-event data [28, 29]. The architecture used for survival modeling mirrored that of the classification task, consisting of four hidden layers with dropout regularization.

For model training, we used 10-fold cross-validation on 210 patients from the full cohort (Cohort I, *n* = 236), while 26 patients were held out as an independent test set for final evaluation. For the Moffitt-only subset (Cohort II, *n* = 131), survival models were trained using 7-fold cross-validation on 105 patients, with 26 patients similarly held out as a test set. Hyperparameters, including the number of nodes per layer, dropout rate, learning rate, weight decay, and random seed, were optimized using Bayesian search via Weights & Biases. Model predictions were averaged across validation folds to generate final outputs.

To enhance robustness and minimize variance, we trained 100 MLP models for each cohort, varying hyperparameters and random seeds. Model performance was evaluated on the held-out test set using the concordance index (C-index), a standard metric for evaluating the discriminatory ability of survival models [30, 31]. Final C-index values represent the mean across all 100 trained models, with 95% confidence intervals’ –distribution. Statistical significance was assessed using paired-sample *t*-tests, comparing each configuration to its immediate predecessor. This ensemble-based strategy enabled a robust evaluation of the prognostic utility of multimodal features, allowing direct comparison as additional data modalities were integrated stepwise (**Figure 1** and **Table 3**).

#### 2.3.5. Analysis of the Performance of LLMs

Three large language models (LLMs) were evaluated for their ability to extract cancer cachexia-related information from unstructured clinical notes: Llama3.2:3b-instruct-fp16, Qwen2.5:7b-instruct-fp16, and DeepSeek-r1:70b. A predefined set of 26 questions pertaining to cachexia symptoms (e.g., weight loss, reduced food intake, muscle wasting) was prepared. Each question was formulated to elicit a categorical response of “yes” “no” or “not given” based on the information present in the clinical notes.

For evaluation, a random sample of 10 patients was selected out of 105 patients with clinical notes. For each of these 10 patients, responses to all 26 questions were manually curated by thoroughly reviewing their unstructured clinical notes to establish ground truth labels. The same set of questions, accompanied by instructions to provide categorical responses along with reasoning and reference, was included in the prompt given to each LLM for inference. The output generated by each LLM was then compared against the manually curated responses to determine the accuracy score. The accuracy score was calculated as the proportion of correctly predicted responses out of the total number of questions per patient.

## 3. Results

We evaluated the performance of our multimodal AI framework across multiple experiments designed to assess the additive value of each data modality for cancer cachexia detection and survival prediction. Results are presented for both the whole cohort (Cohort I, n = 236) and the Moffitt-only cohort (Cohort II, n = 131), following the stepwise modeling configurations described in **Figure 1** and **Table 3**. Each experiment was analyzed for performance using metrics of classification accuracy, precision, recall, F1 score, and concordance index for survival prediction. In addition, we conducted detailed sample-level error analysis, model confidence evaluation, and LLM comparison for clinical note feature extraction.

### 3.1 Model Performance Across Data Modalities

#### 3.1.1. Binary Cachexia Classification Performance

**Table 4** summarizes the binary cancer cachexia classification performance across different combinations of data modalities for both patient cohorts. Consistent with the stepwise modeling framework outlined in **Figure 1**, each experimental configuration, defined in **Table 3**, added a new data source to assess its incremental impact on model performance.

**Table 4:** Binary cancer cachexia classification performance across stepwise data modality configurations for both patient cohorts.

| Performance Metrics | Input Data Modality Configurations |  |  |  |  |  |
| --- | --- | --- | --- | --- | --- | --- |
|  | Clinical + SM | + Lab | + Notes | - SM | + Notes Embeddings | + CT Embeddings |
| **Cohort I ($n = 236$)** |  |  |  |  |  |  |
| Accuracy | 0.69 | 0.73 | **0.85** | 0.81 | 0.88 | **0.92** |
| Precision | 0.65 | 0.69 | **0.80** | 0.75 | **0.92** | 0.86 |
| Recall | 0.85 | 0.85 | **0.92** | 0.92 | 0.85 | **1.00** |
| F1 score | 0.73 | 0.76 | **0.86** | 0.83 | 0.88 | **0.92** |
| **Cohort II ($n = 131$)** |  |  |  |  |  |  |
| Accuracy | 0.77 | 0.81 | **0.85** | 0.81 | 0.77 | **0.81** |
| Precision | 0.73 | 0.79 | **0.85** | 0.79 | 0.77 | **0.83** |
| Recall | **0.85** | **0.85** | **0.85** | **0.85** | **0.77** | **0.77** |
| F1 score | 0.79 | 0.81 | **0.85** | 0.81 | 0.77 | **0.80** |

\* Column details: The configurations in the table (left to right columns) represent the stepwise integration of data modalities, as illustrated in **Figure 1** and defined in **Table 3**. The highest value in each row is shown in **bold**. Rows are grouped into two categories: non-embedding and embedding-based modality integration.
Clinical + SM: Clinical data (sex, race, ethnicity, age, weight, height, BMI, cancer stage), SMA, SMI, SM-HU.
- + Labs: Clinical data, SMA, SMI, SM-HU, serum albumin, neutrophil count, lymphocyte count, blood-urea-nitrogen, serum creatinine, NLR, UCR, CXI, mCXI. - + Notes: Clinical data, SMA, SMI, SM-HU, serum albumin, neutrophil count, lymphocyte count, blood-urea-nitrogen, serum creatinine, NLR, UCR, CXI, mCXI, structured features extracted from unstructured clinical notes using LLMs. - -SM: Clinical data, serum albumin, neutrophil count, lymphocyte count, blood-urea-nitrogen, serum creatinine, NLR, UCR, CXI, mCXI, structured features extracted from clinical notes using LLMs. - + Note Embedding: Clinical data, SMA, SMI, SM-HU, serum albumin, neutrophil count, lymphocyte count, blood-urea-nitrogen, serum creatinine, NLR, UCR, CXI, mCXI, embeddings generated from focused clinical text pertaining to cancer cachexia symptoms. - + CT Embedding: Clinical data, SMA, SMI, SM-HU, serum albumin, neutrophil count, lymphocyte count, blood-urea-nitrogen, serum creatinine, NLR, UCR, CXI, mCXI, focused clinical text embeddings, CT image embeddings from the L3 vertebral level.

For Cohort I (*n* = 236), the baseline model (Clinical + SM) trained using only clinical variables and skeletal muscle metrics achieved an accuracy of 69%, with a precision of 65% and an F1 score of 73%. Adding laboratory data and derived cachexia indices (+ Lab) improved performance across all metrics, increasing accuracy to 73%, precision to 69%, and F1 score to 76%. Further inclusion of structured symptom features (+ Notes) extracted from unstructured clinical notes using LLM resulted in a substantial performance gain, with the model achieving 85% accuracy, 80% precision, and an F1 score of 86%.

When skeletal muscle metrics were removed (–SM) from the inputs while retaining clinical data, laboratory data, and structured note features, accuracy dropped slightly to 81%, with a corresponding decrease in precision and F1 score. This highlights the importance of radiologic muscle metrics for cachexia detection.

Embedding-based models (last two columns in **Table 4**) yielded the highest classification performance. When embeddings of tabular data (clinical variables, skeletal muscle metrics, and laboratory data) were concatenated with GatorTron-generated embeddings from focused clinical text (+ Notes Embeddings), the model achieved an accuracy of 88%, with a precision of 92% and an F1 score of 88%. Adding CT image embeddings (+ CT Embeddings) from the L3 vertebral level further improved accuracy to 92%, with a final precision of 86% and an F1 score of 92%.

Similar trends were observed in Cohot II (*n* = 131). The baseline model integrating clinical data and skeletal muscle metrics achieved 77% accuracy, which improved to 81% with the addition of laboratory data and 85% with the inclusion of structured clinical notes. Excluding skeletal muscle measurements reduced the accuracy back to 81%. Embedding-based models achieved accuracies of 77% and 81% for the two multimodal configurations, with F1 scores of 77% and 80%.

Overall, each stepwise addition of data modalities led to measurable improvements in cachexia classification performance. Embedding-based models that integrated structured, unstructured, and imaging data achieved the highest accuracy and F1 scores across both cohorts.

#### 3.1.2. Patient-Level Prediction Patterns and Error Analysis

To better understand how different data modalities influenced individual patient predictions, we conducted a patient-level error analysis across the stepwise experimental configurations. **Figure 3** and **Figure 4** illustrate correct and incorrect cachexia predictions for each of the 26 test set patients in Cohorts I and II, respectively.

**Figure 3:**
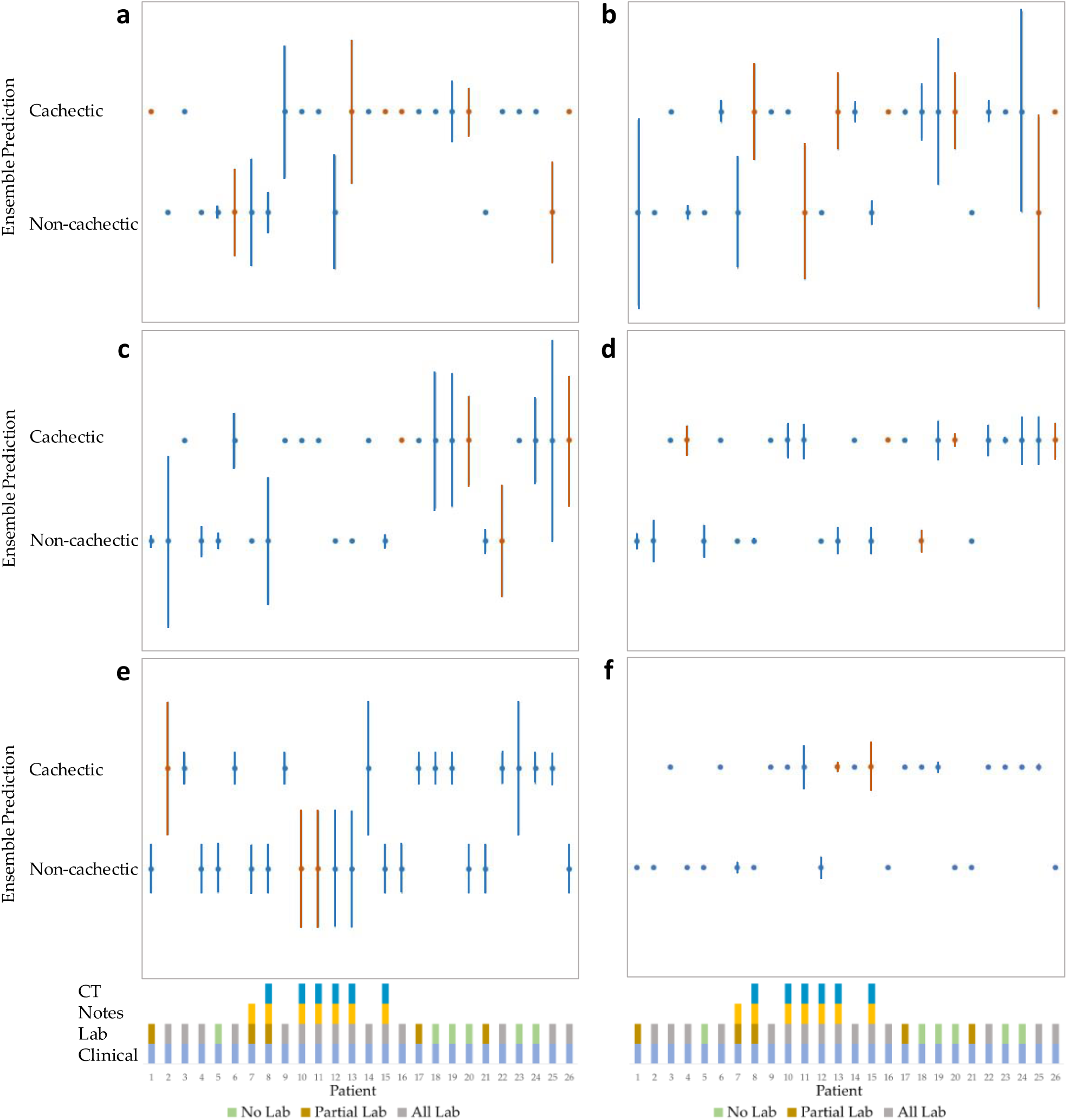
Patient-wise model predictions and confidence estimates for 26 test set patients from Cohort I. Each subfigure (a–f) corresponds to a stepwise data modality configuration as defined in **Table 3**. The x-axis denotes individual patients, and the y-axis shows the predicted cachexia status from an ensemble of five independently trained models. Vertical lines indicate prediction uncertainty; longer bars reflect lower model confidence. Correct predictions are shown in blue, and incorrect predictions in red. The bottom row summarizes available data modalities per patient, with lab data categorized as “No Lab,” “Partial Lab,” or “All Lab” Higher uncertainty is often observed in patients where available data modalities offered limited or conflicting evidence, providing context for prediction reliability across modality configurations.

**Figure 4:**
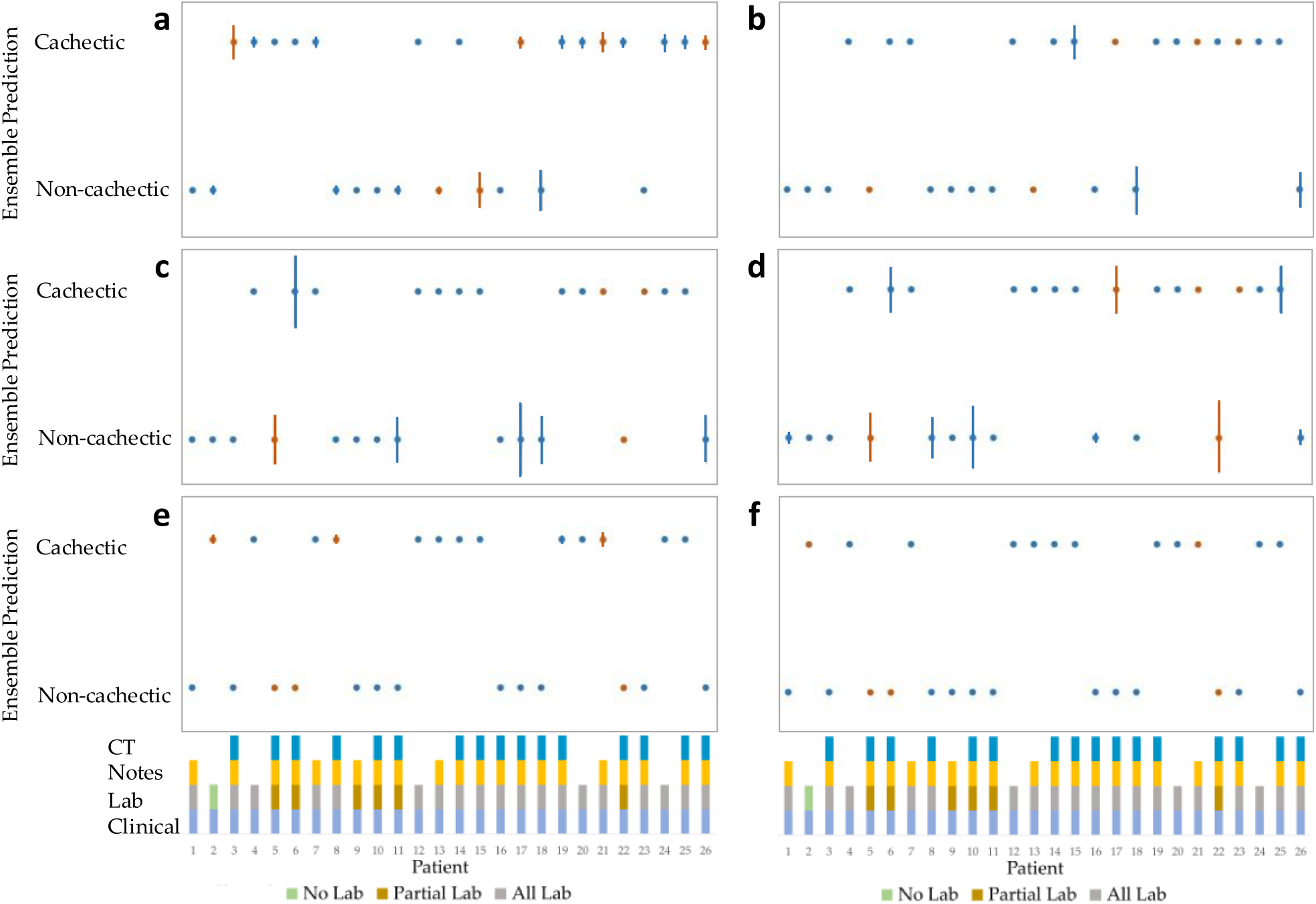
Patient-wise model predictions and confidence estimates for 26 test set patients from Cohort II. Each subfigure (a–f) corresponds to a stepwise data modality configuration as defined in **Table 3**. The x-axis denotes individual patients, and the y-axis shows the predicted cachexia status from an ensemble of five independently trained models. Vertical lines indicate prediction uncertainty; longer bars reflect lower model confidence. Correct predictions are shown in blue, and incorrect predictions in red. The bottom row summarizes available data modalities per patient, with lab data categorized as “No Lab,” “Partial Lab,” or “All Lab” Higher uncertainty is often observed in patients where available data modalities offered limited or conflicting evidence, providing context for prediction reliability across modality configurations.

In Cohort I (**Figure 3**), several patterns emerged. For example, in the baseline model trained with clinical data and skeletal muscle metrics (**Figure 3a**), patients 1, 6, and 15 were misclassified despite having SMI values suggestive of cachexia. When laboratory data was added (**Figure 3b**), these patients were correctly classified, demonstrating the additive diagnostic value of inflammatory and nutritional biomarkers. However, for patients 8 and 11, who were correctly classified using SMI alone, the addition of laboratory data led to misclassification, likely due to discordance between SMI and lab-based cachexia indicators. The inclusion of structured clinical note features (**Figure 3c**) corrected these errors by providing additional context regarding functional status and weight history, as documented in the EHR.

Similar patterns were observed in more complex cases. For example, patient 13 was misclassified by the model trained with clinical, skeletal muscle metrics, and laboratory data, as all these inputs supported an incorrect cachexia status. However, the addition of clinical notes corrected this error, suggesting that text-based information contained compensatory signals not captured in the rest of the data modalities. In contrast, patient 25 lacked clinical notes but was still correctly classified by the model trained with note features, suggesting the presence of indirect signals learned from other patients during training.

The absence of skeletal muscle metrics negatively affected model performance in some cases. For example, patient 4 was misclassified when skeletal muscle metrics were excluded (**Figure 3d**), as these metrics had previously provided strong evidence for cachexia status. In embedding-based models (**Figure 4e** and **Figure 4f**), which integrated all data modalities and were followed by the addition of embeddings from the CT images, improved overall classification accuracy and confidence, particularly for patients with missing or conflicting information in individual modalities. Notably, patients with discordant skeletal muscle metrics and laboratory data (e.g., patients 22 and 26) were correctly classified only after full multimodal embedding was applied. Similar findings were observed in Cohort II (**Figure 4**). Incremental addition of laboratory data and clinical note features improved classification performance, especially in cases with ambiguous skeletal muscle metrics. Embedding-based models further enhanced performance and reduced uncertainty, especially for patients with incomplete data. This error analysis highlights the complementary value of each data modality and supports the importance of multimodal integration for robust cachexia detection.

### 3.2 Model Confidence Analysis

#### 3.2.1 Model Trained Using Clinical + SM (Config 1)

In **Figure 3a**, the model’s confidence largely aligns with the available skeletal muscle metrics (SMI, SM-HU) and the actual cachexia status, with a few notable exceptions. Patient 1 was misclassified as non-cachectic with high confidence. This is because SMI was below the cutoff, and SM-HU was well below the normal range, which strongly supported the incorrect prediction. Patients 2, 3, 4, and 5 were correctly classified, with high confidence, as their SMI values reinforced the model’s predictions. Patient 6 was incorrectly classified with low confidence, as its SMI coincided with the cutoff value, making the prediction uncertain. Similarly, patients 7, 8, and 9 were correctly classified, but their SMI values were close to the cutoff, leading to lower model confidence. Patient 12 was predicted as non-cachectic due to skeletal muscle metrics, CXI, and UCR supporting this status. However, a high neutrophil count (indicative of cachexia) and an unusually high lymphocyte count (not indicative of cachexia) caused the model to be underconfident. Patients 15, 16, and 25 had skeletal muscle metrics that did not indicate cachexia, yet the actual status was cachectic. This led to confident incorrect predictions for patients 15 and 16 and an underconfident incorrect prediction for patient 25. The remaining confident and correct predictions were primarily supported by the skeletal muscle metrics, with a few exceptions. In **Figure 4a**, for most of the predictions, the model output shows high confidence. Patients 3, 15, and 18 have slightly low confidence, with the SM-HU and SMI having conflicting values in patient 3 and the BMI and SMI having conflicting values in patient 15. However, in patient 18, the BMI and SMI support the non-cachectic status, which is correct.

#### 3.2.2 Model Trained Using Clinical + SM + Lab (Config 2)

In **Figure 3b**, the model’s confidence was influenced by conflicting information between skeletal muscle metrics and laboratory data. Patient 1 was correctly classified as cachectic, but with low confidence, due to conflicting SMI and lab data values. Patients 2, 3, and 4 also had conflicting skeletal muscle metrics and lab data values, but the model maintained high confidence in its correct predictions. Patient 5 had missing lab data, yet the model remained confident in its prediction based on skeletal muscle metrics alone. Patient 6 was incorrectly classified, with all included modalities supporting the incorrect decision; however, an unusually high lymphocyte count (unrelated to cachexia) seemed to influence a confident, incorrect prediction. Patients 7 and 8 had skeletal muscle metrics at the cut-off point, and patient 7 also had lab data values near the normal range, leading to a correct but low-confidence prediction. Patient 8 had a serum creatinine level slightly below the normal (not indicative of cachexia), but the model used this abnormality to make an incorrect decision with low confidence. Patients 9 and 10 were correctly classified as cachectic, as SMI and high neutrophil counts supported this status. However, CXI was high (suggesting non-cachexia), due to elevated lymphocyte counts. The model likely relied more on neutrophil levels for a confident, correct prediction. Patients 11, 13, 20, and 25 had conflicting skeletal muscle metrics and lab data values, leading to low-confidence predictions. Patients 16 and 26 were incorrectly classified with high confidence because patient 16 had a high neutrophil count and low skeletal muscle metrics that strongly supported the incorrect decision. Patient 26 had low skeletal muscle metrics, high UCR, and low BUN, suggesting muscle loss and malnutrition, leading to a confident incorrect prediction. Patient 19 was correctly classified as non-cachectic, but with low confidence, since skeletal muscle metrics contradicted the actual status. Patient 24 had skeletal muscle metrics supporting cachexia, but the model trained with the addition of lab data made a correct prediction with low confidence due to missing lab data for this patient. In **Figure 4b**, patients 15, 18, and 26 are underconfident because the lab data, BMI, and SMI have conflicting information.

#### 3.2.3 Model Trained Using Clinical + SM + Lab + Notes (Config 3)

In **Figure 3c**, model confidence was influenced by the availability and content of clinical notes. Patients 2, 8, 18, 19, and 25 were correctly classified, but with low confidence, as no clinical notes were available to reinforce the prediction. Patient 8 had notes that partially supported the correct non-cachectic status, such as unintentional weight loss after surgery, a normal Karnofsky score (assessing the patient’s physical performance), and loss of appetite. Patients 1, 3, 4, 5, 6, 9, 14, 17, 21, and 23 had no available notes but maintained confident predictions based on prior knowledge from lab data, except for patient 1, where the model trained with addition of lab data made a low-confidence decision, but the model trained with the addition of notes was confident. Patients 7, 10, 11, 12, 13, and 15 had clinical notes supporting the actual status, leading to correct and confident predictions, except for patient 13, where the notes partially support cachectic status, but this was not the actual status. Patient 16 remained incorrectly classified, as no clinical notes were available. **Figure 4c** shows low confidence for patients 5, 6, 11, 17, 18, and 26. The notes for patients 5, 11, 17, 18, and 26 support the correct cachexia status. However, some symptoms extracted from the notes conflict with the correct status, causing low confidence. In the case of patient 6, the notes support the incorrect non-cachectic status, although the model makes the correct prediction, which explains the low confidence of the model in its prediction.

#### 3.2.4 Model Trained Using Clinical + Lab Data + Notes (No SM) (Config 4)

In **Figure 3d**, the model made confident decisions. For patients 2 and 25, the conflicting information between different lab values made the model a little underconfident. For patients 19 and 24, there are no additional modalities; therefore, when skeletal muscle metrics are removed, the model is underconfident. **Figure 4d** presents patients 5, 6, 8, 10, 17, and 22 to be underconfident when skeletal muscle metrics are not used to train the model. Patients 5, 6, and 17 have relatively low confidence because of some conflicting symptoms in the notes. Notes for patients 8 and 10 support the correct status, with lab data and BMI also supporting the correct status. However, there are a few symptoms, such as weight loss after surgery and early satiety, that cause the model to become underconfident. Notes for patient 22 support a non-cachectic status that is incorrect and conflicts with the BMI and partial reports when skeletal muscle metrics supporting the correct status are removed.

#### 3.2.5 Model Trained Using Clinical + SM + Lab + Notes Embeddings (Config 5)

In **Figure 3e**, model confidence was influenced by conflicting information across multiple modalities. Patients 2, 10, 11, 12, 13, 14, and 23 were predicted with low confidence due to conflicting information from different data sources. Patients 14 and 23 were exceptions, where data correctly supported the actual cachexia status, but the model remained underconfident. In **Figure 4e**, patients 11, 12, and 15 are underconfident, with all data modalities for patient 11 supporting the correct cachexia status. However, the notes had some symptoms that can cause the model to become underconfident, such as weight loss. Patient’, but the skeletal muscle metrics and the lab data have conflicting information. Patient 15 has conflicting lab data and notes with symptoms partially supporting the correct cachectic status.

**Figure 5:**
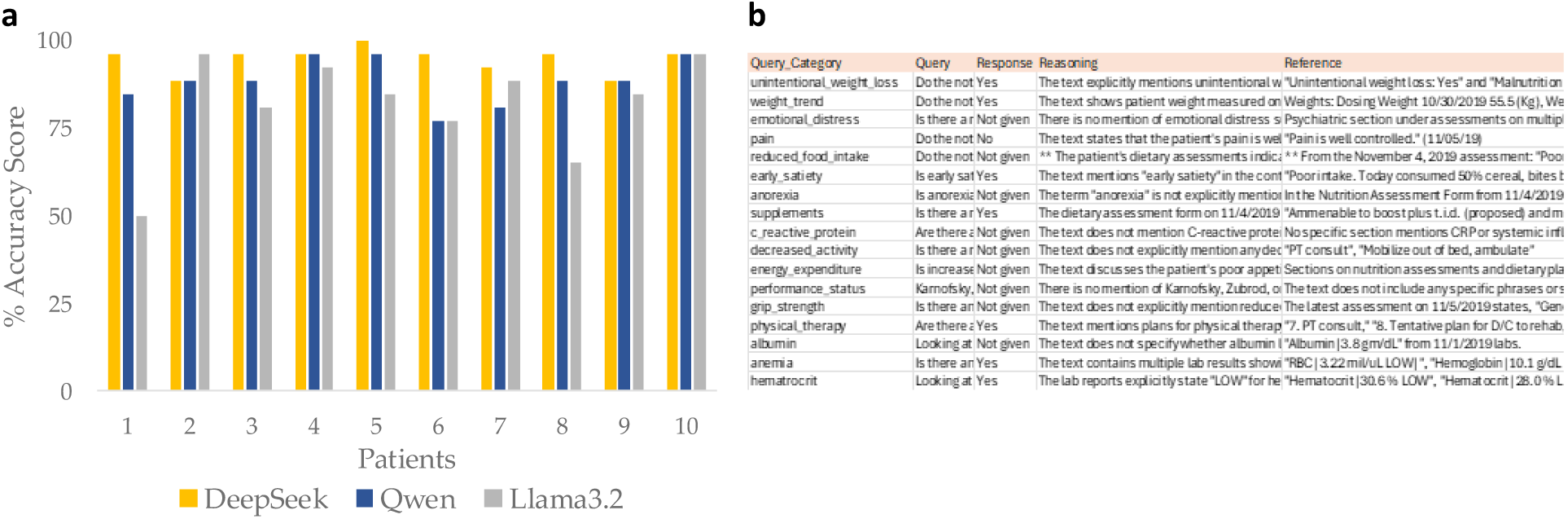
(**a**) Performance comparison of the three LLMs evaluated for extracting structured data from unstructured clinical notes, with DeepSeek achieving the highest average accuracy score, followed by Qwen. The accuracy score per patient was calculated as the percentage of questions correctly answered based on information extracted from each patient’s’clinical notes. Performance was assessed in terms of average accuracy score across ten randomly selected representative patients to compare the LLMs. (**b**) Example of the tabular output template generated by the LLMs, showing a subset of the input questions and their corresponding responses, including reasoning and references from the unstructured clinical notes for a single patient.

#### 3.2.6 Model Trained Using Clinical + SM + Lab + Notes + CT Embeddings (Config 6)

In **Figure 3f**, model decisions are influenced by information from CT images. In **Figure 4f**, all predictions are confident in terms of variance between the ensemble models. This analysis considers additional data modalities (skeletal muscle metrics, laboratory data, and clinical notes) rather than the clinical data, which also impacted the model’s overall confidence in its decision.

### 3.3 Survival Analysis using Various Modalities of Data

**Table 5** presents the comparison of the concordance index (C-Index) across different combinations of data modalities. When using only clinical data, the C-Index on the test set is 0.727 for Cohort I and 0.638 for Cohort II. Adding skeletal muscle metrics (SMA, SMI, and SM-HU) improves the C-Index to 0.743 and 0.656, respectively, indicating the predictive value of body composition features. In Cohort I, the addition of lab data results in a drop in C-Index to 0.663, likely due to missing values across the additional modality. In contrast, the C-Index improves to 0.670 in Cohort II with the addition of lab data, benefiting from a lower difference in missingness between the training and testing sets. In the training set, lab data were available for 71.0% patients in Cohort I and 73.3% in Cohort II. Within the test set, the availability of lab data was 61.5% in Cohort I and 73.1% in Cohort II. Adding clinical notes further improves the C-Index to 0.671 in Cohort I and 0.722 in Cohort II. The greater improvement in Cohort II correlates with higher availability of notes in the test set (61.5% vs. 23.1% in Cohort I). Finally, when all other modalities are included but skeletal muscle metrics are removed, the C-Index drops to 0.659 in Cohort I and 0.713 in Cohort II, confirming the contribution of skeletal muscle metrics to model performance. All changes in C-index across modality combinations were statistically significant (*p* < 0.05), as determined by paired-sample t-tests comparing results from 100 independently trained models evaluated on the same test set.

**Table 5:**
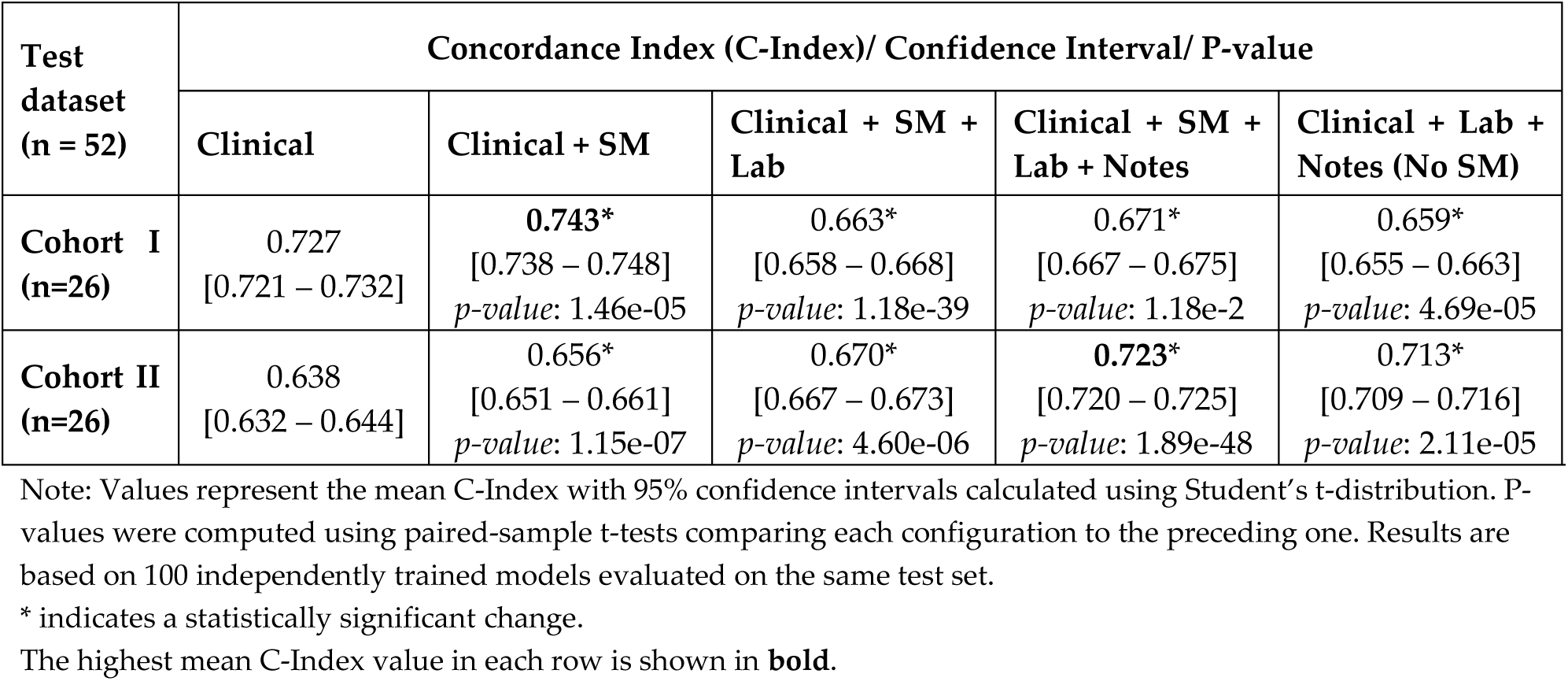
Mean Concordance Index (C-Index) and 95% Confidence Intervals (CIs) across data modality combinations.

### 3.4 Performance Comparison of LLMs for Extracting Structured Data from Unstructured Clinical Notes

As presented in **Figure 5**, DeepSeek achieved the highest average accuracy score of 94.62% across the ten randomly selected patients, demonstrating its effectiveness in transforming unstructured clinical notes into structured cachexia features. Qwen followed with an average accuracy score of 88.46%, while Llama3.2 achieved an average accuracy score of 81.54%. Notably, all three models performed similarly for patients 4, 9, and 10.

## 4. Discussion

Cancer cachexia remains a major clinical challenge due to its heterogeneous and multifactorial nature, encompassing systemic inflammation, metabolic dysfunction, muscle wasting, and nutritional decline [32, 33]. Commonly used indicators, including anthropometric changes, skeletal muscle index, and laboratory biomarkers such as C-reactive protein (CRP), offer only partial insights and lack specificity. These markers are not independently diagnostic of cancer cachexia, as they may also reflect other underlying conditions [3, 32, 34]. Accurate diagnosis requires integrating information across multiple domains: skeletal muscle depletion visible on imaging, metabolic and inflammatory abnormalities from laboratory tests, and functional and symptomatic changes documented in clinical notes. Composite indices such as CXI, mCXI, and CCRS aim to unify these dimensions but rely on rigid thresholds specific to cancer types, limiting their adaptability in diverse patient populations and routine clinical workflows [4–8].

To address these limitations, we developed and evaluated a multimodal, uncertainty-aware AI framework for early detection of cancer cachexia using routinely collected, real-world clinical data. The framework integrates structured clinical and laboratory variables, imaging-derived skeletal muscle metrics, unstructured clinical notes, and embeddings from radiological scans. Its modular design allows flexible integration of available data modalities while tolerating missing inputs, supporting scalability across diverse clinical settings. By generating individualized, patient-specific predictions and quantifying predictive uncertainty, the framework enables human-in-the-loop clinical decision-making and supports safe, interpretable deployment within oncology workflows.

A key innovation of this study is the framework’s comprehensive multimodal design, which incrementally integrates heterogeneous data modalities reflecting the complex pathophysiology of cancer cachexia. Starting with structured clinical variables and skeletal muscle metrics, model performance improved progressively with the addition of laboratory biomarkers, structured symptom features extracted from unstructured clinical notes using LLMs, and radiology-derived imaging embeddings. By leveraging LLMs, the framework transformed free-text clinical documentation, including nutritional assessments, functional status reports, and symptom narratives, into structured features that substantially enhanced prediction performance. Embedding-based models unified high-dimensional representations across modalities, resulting in the highest observed accuracy, precision, and F1 scores. Beyond classification performance, the framework’s uncertainty-aware design represents a critical contribution. As additional modalities were integrated, prediction uncertainty decreased systematically, supporting safer and more reliable clinical deployment. By quantifying predictive confidence and flagging low-confidence cases, the system enables human-in-the-loop oversight, ensuring that ambiguous cases are referred to clinical experts, an essential safeguard for ethical AI implementation. Moreover, the framework is engineered to accommodate missing modalities without requiring data imputation or case exclusion, supporting scalability and reducing exclusion bias across diverse clinical environments.

From a clinical integration standpoint, this framework addresses key barriers to the adoption of AI tools for cachexia detection. By leveraging routinely collected data across clinical, laboratory, imaging, and unstructured note modalities, and by accommodating incomplete records, it reflects the realities of oncology workflows and diverse clinical environments. The uncertainty-aware design reduces automation bias and supports human-in-the-loop decision-making by flagging low-confidence cases for clinician review, promoting safety and interpretability. Unlike static threshold-based indices such as CXI or CCRS, which are limited by cancer-type-specific cutoffs, our patient-specific modeling approach enhances fairness, adaptability, and equitable performance across heterogeneous patient populations. The framework’s modular architecture and local deployability also support integration within EMR systems or oncology decision-support platforms, aligning with broader goals in medical AI to deliver scalable, interpretable, and clinically actionable tools for precision oncology.

While these findings are promising, several limitations warrant consideration and guide important directions for future work. First, the study focused exclusively on patients with PDAC and was conducted within a single statewide research consortium, with Moffitt Cancer Center contributing the imaging and clinical note data for this study. As illustrated in **Figure 1**, our stepwise, modular framework was explicitly designed to accommodate real-world variability in data availability; however, the cohort distributions (**Figure 2**) highlight patterns of missingness that may affect generalizability. Second, although our use of LLMs for structured feature extraction from clinical notes enhanced model performance, variability in documentation practices across institutions may limit reproducibility, underscoring the need for external validation. Third, the radiologic skeletal muscle metrics were derived from a single anatomical level (L3) and imaging phase, which may reduce applicability to other imaging protocols or anatomical landmarks.

Future work will focus on expanding and validating the framework across multi-institutional, multi-cancer datasets and diverse care settings. Incorporating longitudinal data, serial imaging, repeated laboratory values, evolving symptom profiles from clinical notes, and novel biomarkers could enhance cachexia monitoring and enable earlier intervention. Moreover, prospective clinical trials, clinician-in-the-loop evaluations, and real-world deployment studies will be essential to assess the framework’s safety, clinical utility, and impact on both cancer cachexia detection and survival prediction. Finally, future deployments should incorporate fairness audits and bias monitoring strategies to ensure equitable model performance across patient subgroups, supporting ethically responsible use of AI in oncology care.

## 5. Conclusion

This study demonstrates the feasibility and clinical utility of a multimodal, uncertainty-aware AI framework for early detection and prognostic assessment of cancer cachexia at the time of diagnosis. By integrating structured clinical variables, laboratory biomarkers, radiologic muscle metrics, unstructured clinical text, and imaging-derived embeddings, the framework delivers improved predictive and prognostic performance while remaining robust to missing data. Leveraging LLMs and foundation models enables scalable feature extraction across diverse data types, supporting generalizability across oncology settings. The incorporation of uncertainty estimation facilitates human-in-the-loop oversight, enhancing clinical safety and trust. This work represents an important step toward real-world deployment of equitable, interpretable, and trustworthy AI-driven decision support for cancer cachexia management and personalized oncology care.

## Author contributions

All authors have read and agreed to this version of the manuscript.

S.A. and G.R. designed the concept and the methodology; S.A. wrote the original draft of the manuscript, prepared the figures, and wrote the software for the framework; S.A., N.P., M.P., E.D., D.J, J.P., M.S., Y.Y., and G.R. validated the study and the results and substantially contributed to shaping the final manuscript; N.P., M.P., J.P., and G.R. provided the data, hardware, and software resources to complete this work.

## Acknowledgements

This study was funded by NSF grants 2234468 and 2234836, NIH grant U01CA200464, James and Esther King Biomedical Research Grant 8JK02, Department of Defense grant W81XWH-22-1-1021, and was funded in part by the Quantitative Imaging Core and the Biostatistics and the Bioinformatics Shared Resource at the H. Lee Moffitt Cancer Center & Research Institute, an NCI designated Comprehensive Cancer Center (P30-CA076292). The funders played no role in study design, data collection, analysis and interpretation of data, or the writing of this manuscript.

## Data Availability

The data supporting this study’s findings are available from the Florida Pancreas Collaborative, but restrictions apply. These data were used under license for the current study and are not publicly available. However, data is available from the authors upon reasonable request and with permission of the Florida Pancreas Collaborative.

## Code Availability

The underlying code for this study is available in GitHub repository *Multimodal AI Biomarker* and can be accessed via this link https://github.com/Beemd/multimodal_AI_biomarker.

## Institutional Review Board Statement

The study was conducted in accordance with the Declaration of Helsinki and approved by the Institutional Review Board (or Ethics Committee) of Moffitt Cancer Center MCC 22299, MCC 19717 (version 1.4 approved on 02/12/2019), and MCC21962.

## Informed Consent Statement

Informed consent was obtained from all subjects involved in the study.

